# Cumulative Febrile, Respiratory, and Diarrheal Illness among Infants in Rural Guatemala and their Association with Neurodevelopmental and Growth Outcomes

**DOI:** 10.1101/2022.09.02.22279419

**Authors:** Daniel Olson, Molly M. Lamb, Amy K. Connery, Alison M. Colbert, Mirella Calvimontes, Desiree Bauer, M. Alejandra Paniagua-Avila, María Alejandra Martínez, Paola Arroyave, Sara Hernandez, Kathryn L. Colborn, Yannik Roell, Jesse J. Waggoner, Muktha S. Natrajan, Evan J. Anderson, Guillermo A. Bolaños, Hana M. El Sahly, Flor M. Munoz, Edwin J. Asturias

**Affiliations:** Department of Pediatrics, University of Colorado School of Medicine, 13123 E. 16th Ave., Aurora, CO 80045, USA; Center for Global Health, Colorado School of Public Health, 13199 East Montview Blvd, Aurora, CO 80045, USA; Children’s Hospital Colorado, 13123 E. 16th Ave., Aurora, CO 80045, USA; Department of Epidemiology, Colorado School of Public Health, 13001 E 17th Pl, Aurora, CO 80045, USA; Department of Physical Medicine and Rehabilitation, University of Colorado School of Medicine, 12631 E 17th Ave, Aurora, CO 80045, USA; Center for Human Development, Fundacion para la Salud Integral de los Guatemaltecos, Retalhuleu, Guatemala; Department of Epidemiology, Columbia University Mailman School of Public Health, 722 West 168th St. New York, NY 10032, USA; Department of Surgery, University of Colorado School of Medicine, 12631 E 17th Ave #6117, Aurora, CO 80045, USA; Department of Medicine, Emory University School of Medicine, 1364 Clifton Road NE, Atlanta, GA 30322, USA; Department of Pediatrics, Emory University School of Medicine, 2015 Uppergate Drive, Atlanta, GA 30322, USA; Department of Molecular Virology and Microbiology; Department of Pediatrics, Baylor College of Medicine, 1 Baylor Plaza, Houston, TX 77030, USA

**Keywords:** fever, cough, neurodevelopment, Guatemala, Mullen Scale of Early Learning (MSEL)

## Abstract

**Objective:** We aimed to evaluate the association between cumulative illness with neurodevelopment and growth outcomes in a birth cohort of Guatemalan infants.

**Study Design:** From June 2017 to July 2018, infants 0-3 months of age living in a resource-limited region of rural southwest Guatemala were enrolled and completed weekly at-home surveillance for caregiver-reported cough, fever and vomiting/diarrhea. They also underwent anthropometric assessments and neurodevelopmental testing with the Mullen Scales of Early Learning (MSEL) at enrollment, six months, and one year.

**Results:** Out of 499 enrolled infants, 430 (86.2%) completed all study procedures and were included in the analysis. At 12-15 months of age, 140 (32.6%) infants had stunting (length-for-age Z [LAZ] score <-2 SD) and 72 (16.7%) had microcephaly (occipital-frontal circumference [OFC] <-2 SD of the mean). In multivariable analysis, greater cumulative weeks of reported cough illness (beta=-0.08/illness-week, p=0.06) and febrile illness (beta=-0.36/illness-week, p<0.001) were marginally or significantly associated with lower MSEL Early Learning Composite (ELC) Score at 12-15 months, respectively; there was no association with any illness (cough, fever, and/or vomiting/diarrhea; p=0.27) or with cumulative weeks of diarrheal/vomiting illness alone (p=0.66). No association was shown between cumulative weeks of illness and stunting or microcephaly at 12-15 months.

**Conclusions:** These findings highlight the negative cumulative consequences of frequent febrile and respiratory illness on neurodevelopment during infancy. Future studies should explore the inflammatory profile associated with these syndromic illnesses and their impact on neurodevelopment in the first years of life.

## INTRODUCTION

Among children under 5 years of age living in low- and lower middle–income countries (LMICs), an estimated 43% are at risk for not meeting their neurodevelopmental potential.^1, 2^ Neurodevelopmental problems likely result from the cumulative environmental exposures of living in poverty, including elevated risk of adverse birth outcomes, malnutrition, infectious and non-infectious disease-related inflammation, and psychosocial stressors, among others.^1^

Stunting is another important outcome among children in LMICs, and it shares risk factors with neurodevelopmental delay, such as chronic malnutrition and enteric disease/dysfunction. Recent evidence suggests, however, that despite this overlap, the pathophysiology of each of these two outcomes (neurodevelopment and stunting) is unique and complicated by multiple clinical and environmental factors, and distinct inflammatory phenotypes.^3-7^

Recurrent enteric disease and dysfunction have been tied to both poor neurodevelopment and stunting.^1, 8^ However, the role and pathogenesis of other types of recurrent infectious diseases, such as febrile and respiratory illnesses, is less well understood. A longitudinal birth cohort study from Bangladesh found that recurrent febrile illness and cytokine profiles associated with both enteric and systemic inflammation were associated with neurodevelopmental delays among infants living in informal urban settlements (slums).^5-7, 9^ To our knowledge, this finding has not been replicated in other populations.

Similar to Bangladesh, Guatemalan children suffer from high rates of communicable illness and similar adverse environmental exposures, including the highest prevalence of stunting (49.8%) in the Americas and the sixth highest prevalence of stunting among children under five in the world.^10-14^ We recently completed a longitudinal cohort study (DMID 16-0057) to evaluate the neurodevelopmental outcomes following post-natal Zika virus infection among infants living in rural, southwest Guatemala. The study collected weekly syndromic illness data, providing an opportunity to conduct this secondary analysis exploring the association between caregiver-reported infectious illness symptoms with growth outcomes (i.e., stunting and microcephaly) and neurodevelopment in this community-based infant cohort.

## METHODS

We conducted a secondary analysis of a prospective cohort study of postnatally acquired Zika virus (ZIKV) infection among infants and young children at the University of Colorado-associated Center for Human Development in the rural lowlands of southwest Guatemala. The site encompasses primarily rural communities with approximately 25,000 residents and is located approximately 30 km from the border of Chiapas, Mexico. These communities are monolingual Spanish-speaking. The population suffers from high rates of year-round food insecurity and child undernutrition, diarrheal disease, maternal depression, and maternal and child morbidity and mortality.^15-17^

From June 2017 to July 2018, 499 infants and their mothers were enrolled at 0-3 months of age into the parent study; ZIKV infection status was unknown at the time of enrollment. In this secondary analysis, we included enrolled subjects who had complete outcome data at 12-15 months. Enrollment included collection of baseline demographic, epidemiologic, clinical, anthropometric, and neurodevelopmental data. Mother and father’s literacy (yes/no) and education were self-reported at enrollment, with education being reported on a 4-point scale (mother/father had completed no, primary, secondary, or university/postgraduate education). Households were queried for food insecurity at enrollment using three questions asking if, in the preceeding 4 weeks: 1) the household didn’t have any food due to lack of resources, 2) if anyone went to bed hungry because there was not enough food, or 3) if anyone in the household went a whole day without eating because there was not enough food.^18^ Anthropometric data were collected at enrollment and 3-month intervals on children, and at enrollment and final visit for mothers. Length-for-age z scores (LAZ) were calculated to determine stunting, defined as <2 standard deviations (SDs) below the mean on World Health Organization (WHO)-defined growth curves.^19^ Head circumference was determined by taking the mean of two occipital frontal circumference (OFC) measurements using the Seca 211 Head Circumference Measuring Tape (12-59 cm) following standard operating procedures, and microcephaly was defined as <2 standard deviations below the mean on WHO-defined growth curves.

Infants underwent neurodevelopmental testing by trained Guatemalan psychologists, supervised by bilingual US-based neuropsychologists. Neurodevelopmental testing included the Mullen Scales of Early Learning (MSEL) with rigorous procedures previously described.^20^ The MSEL is comprised of five subscales: Gross and Fine Motor, Expressive and Receptive Language, and Visual Reception. The scores from four of the scales (excluding Gross Motor) are then summed to create the Early Learning Composite Score. The instrument underwent extensive validation and adaptation to the local population at the study site in Guatemala.^21, 22^

Following enrollment, households were visited weekly by trained study nurses, and caregivers were asked to report presence/absence of each of the following symptoms in the preceding week: fever, cough, vomiting/diarrhea (≥3 loose stools/day), rash, conjunctivitis (non-purulent/hyperemic), arthralgia, myalgia, or peri-articular edema. The weekly household visits were continued for 12 months.

Anthropometric data were collected, and medical examination and neurodevelopmental assessments (including the MSEL) were also conducted at 6-9 and 12-15 months of age on all children.

Descriptive statistics were used to characterize the study population. Univariate and multivariable linear and generalized linear regression models were used to test associations between cumulative weeks of each syndromic illness of interest (fever, cough, vomiting/diarrhea, or any of the three illnesses) in infancy (exposures), and 12-15-month MSEL Early Learning Composite (ELC) score (continuous outcome), MSEL subscale scores (continuous outcome), stunting (dichotomous outcome), and microcephaly at 12-15 months (dichotomous outcome). Beta estimates are presented for continuous outcomes, and relative risks (RR) are presented for dichotomous outcomes. After identification of potentially relevant confounders via review of the literature, we conducted a bivariate analysis, adjusted for age at MSEL collection, of the following individual variables for association with MSEL scores, stunting, and microcephaly: sex, maternal height, maternal/paternal literacy, maternal/paternal education, housing material (proxy for socioeconomic status), food insecurity, community (dichotomized into ‘high risk for infectious illness’ vs. ‘not high risk for infectious illness’), and breastfeeding status at study completion. We then selected all variables with a p-value <0.20 in the univariate analysis to be included in the multivariate analysis. In the case of stunting and microcephaly, any variable with a p-value<0.20 for either of these anthropometric outcomes was included in the multivariate analyses for both outcomes. We assessed collinearity of the education variables (maternal/paternal literacy and education) using Spearman correlation coefficients. We chose maternal education as the most relevant variable to include in the multivariable analyses. As enrollment occurred after birth, and gestational age and birthweight are rarely documented in this community (and so retrospective caregiver-report is the only available data), those potential confounders were not considered for these multivariable models. Multivariable models of MSEL outcomes were not adjusted for stunting, which is a separate 12-15-month outcome that shares complex and incompletely overlapping causal pathways with neurodevelopment.^23-28^

All descriptive analyses and regression modeling were conducted using SAS version 9.4 (SAS Institute, Cary, NC) and maps were created in ArcGIS Pro (Esri, Redlands, CA). Global Positioning System (GPS) coordinates of households were used to map cumulative syndromic illness by geolocation. Weeks of cumulative illness for fever, cough, vomiting/diarrhea, and any illness were averaged by community to visualize the spatial distribution of disease burden. The averages for each community were compared to the average of all other communities using the Mann Whitney U Test (p<0.05), to identify communities with significantly greater disease burden (‘high risk for infectious illness’).

The study protocol was reviewed and approved by the Institutional Review Board at Baylor College of Medicine, the Colorado Multiple Institutional Review, the National Ethics Committee of the Guatemalan Ministry of Public Health and the local Community Advisory Board in Trifinio, Guatemala.

## RESULTS

Of 499 enrolled infants in the parent study, 430 (86%) completed the 12-month study and were included in this analysis. The analysis cohort had a mean enrollment age of 1.45 months; 223 (51.9%) were male, 392 (91.4%) mothers self-reported to have functional literacy skills, and 323 (75.5%) were breastfeeding at study completion (**Table 1**). Over the 12 months of observation, infants had caregiver-reported illness for a median of 16 weeks (IQR 10-22 weeks). Cough was reported most frequently (median=11 weeks, IQR 7-17 weeks), followed by diarrhea/vomiting (5 weeks, IQR=3-8 weeks), and fever (3 weeks, IQR=1-5 weeks). Male infants had a non-significant trend towards higher weeks of illness in the first year of life than females (17.3 vs 15.9 weeks, p=0.07).

**Table 1:**
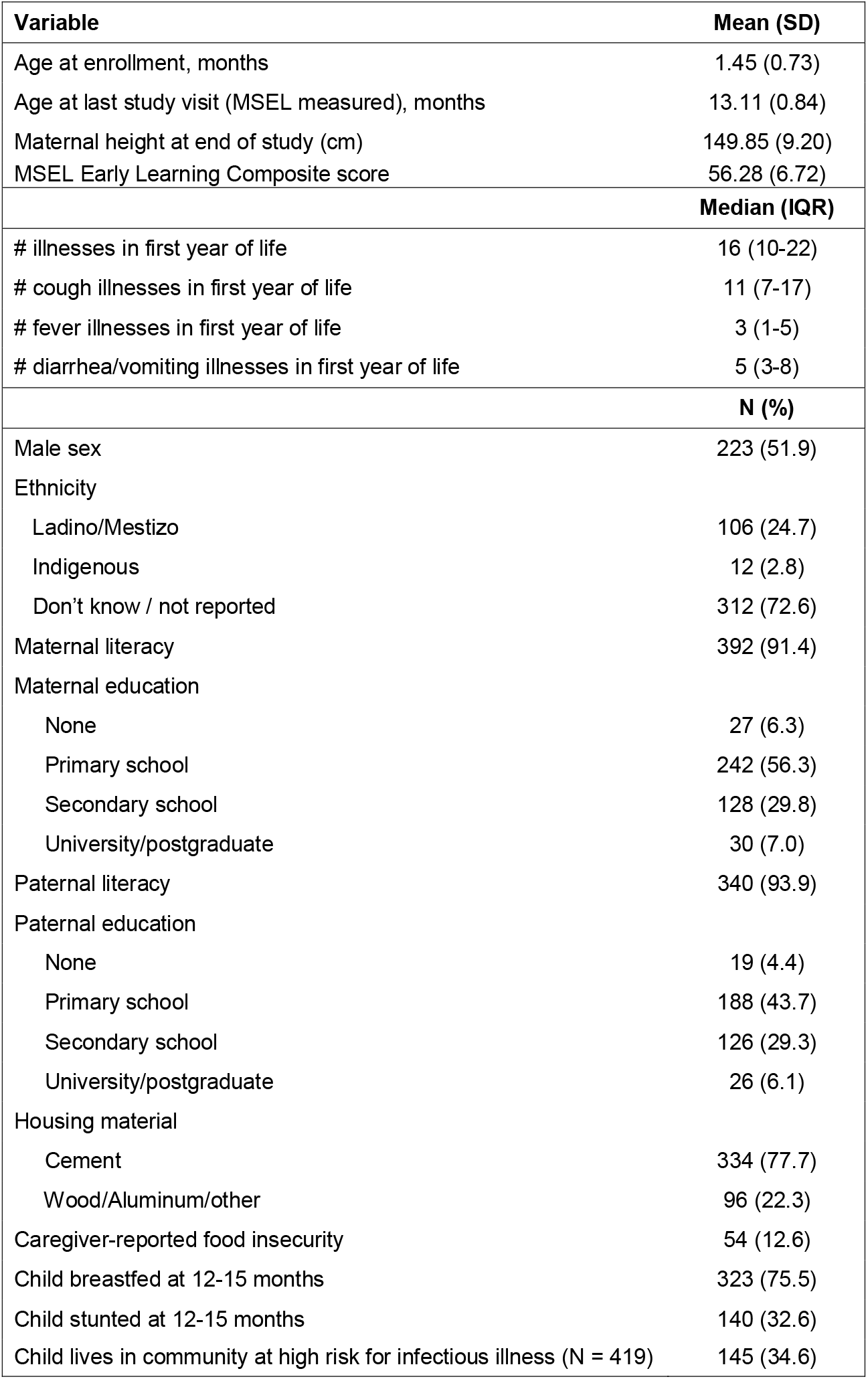
Subject characteristics of infants who completed study follow-up in rural Southwest Guatemala, 2017-19 (n=430).

| **Variable** | **Mean (SD)** |
| --- | --- |
| Age at enrollment, months | 1.45 (0.73) |
| Age at last study visit (MSEL measured), months | 13.11 (0.84) |
| Maternal height at end of study (cm) | 149.85 (9.20) |
| MSEL Early Learning Composite score | 56.28 (6.72) |
|  | **Median (IQR)** |
| # illnesses in first year of life | 16 (10-22) |
| # cough illnesses in first year of life | 11 (7-17) |
| # fever illnesses in first year of life | 3 (1-5) |
| # diarrhea/vomiting illnesses in first year of life | 5 (3-8) |
|  | **N (%)** |
| Male sex | 223 (51.9) |
| Ethnicity |  |
| Ladino/Mestizo | 106 (24.7) |
| Indigenous | 12 (2.8) |
| Don't know / not reported | 312 (72.6) |
| Maternal literacy | 392 (91.4) |
| Maternal education |  |
| None | 27 (6.3) |
| Primary school | 242 (56.3) |
| Secondary school | 128 (29.8) |
| University/postgraduate | 30 (7.0) |
| Paternal literacy | 340 (93.9) |
| Paternal education |  |
| None | 19 (4.4) |
| Primary school | 188 (43.7) |
| Secondary school | 126 (29.3) |
| University/postgraduate | 26 (6.1) |
| Housing material |  |
| Cement | 334 (77.7) |
| Wood/Aluminum/other | 96 (22.3) |
| Caregiver-reported food insecurity | 54 (12.6) |
| Child breastfed at 12-15 months | 323 (75.5) |
| Child stunted at 12-15 months | 140 (32.6) |
| Child lives in community at high risk for infectious illness (N = 419) | 145 (34.6) |

### Neurodevelopment

In univariate regression models to identify potential confounders, higher age-adjusted MSEL ELC Score at 12-15 months was significantly associated with the following variables: maternal height (β=0.23, CI=0.13 – 0.33, p<0.001), maternal literacy (β=3.97 CI=1.90 – 6.05, p<0.001), maternal education (β=2.03, CI=1.22 – 2.85, p<0.001), child not breastfeeding at 12-15 months (β=-1.63, CI=-1.63 --0.28, p=0.02), paternal education (β=0.89, CI=-0.001 – 1.79, p=0.05), and living in a community at high risk for infectious illness (β=-2.44, CI=-3.65 – −1.23, p=<0.0001) (**Table 2**).

**Table 2:** Bivariate* association between demographic and clinical variables and MSEL Early Learning Composite (ELC) Score at 12-15 months. *\*adjusted for age at MSEL collection*

| **Variable** | **Beta estimate (95% Confidence Interval)** | **p-value** |
| --- | --- | --- |
| # illnesses in first year of life | -0.08 (-0.16 - -0.01) | 0.03 |
| # cough illnesses in first year of life | -0.12 (-0.20 - -0.04) | 0.005 |
| # fever illnesses in first year of life | -0.40 (-0.61 - -0.20) | <0.001 |
| # diarrhea/vomiting illnesses in first year of life | -0.05 (-0.17 - 0.07) | 0.41 |
| Male sex | -1.06 (-2.24 - -0.13) | 0.08 |
| Maternal height (cm) at end of study | 0.23 (0.13-0.33) | <0.001 |
| Maternal literacy | 3.97 (1.90 – 6.05) | <0.001 |
| Maternal education | 2.03 (1.22 – 2.85) | <0.001 |
| Paternal literacy | 1.05 (-1.59 – 3.69) | 0.44 |
| Paternal education | 0.89 (-0.001 – 1.79) | 0.050 |
| Housing made of wood/aluminum | -2.13 (-3.54 - -0.72) | 0.003 |
| Food insecurity | -1.49 (-3.27 – 0.30) | 0.10 |
| Child breastfed at 12-15 months | -1.63 (-2.98 - -0.28) | 0.02 |
| Child lives in community at high risk for infectious illness | -2.44 (-3.65 - -1.23) | <0.001 |

After adjusting for age at last visit (12-15 months) and variables identified on univariate analysis with p<0.2, the cumulative number of weeks with reported cough (beta estimate=-0.08 per illness-week, CI=- 0.16 – 0.003, p=0.06) or fever (beta estimate=-0.36 per illness-week, CI=-0.56 - −0.16, p<0.001) had a non-significant trend or were significantly associated with decreased 12-15-month MSEL ELC score (**Table 3**); there was no association with any illness (cough, fever, or vomiting/diarrhea: p = 0.27) or diarrhea/vomiting (p=0.60). Weeks of fever was significantly associated or trended towards significance with all ELC subdomain scores at 12-15 months (p=0.055 to 0.002; Table 3). Gross motor subdomain MSEL score at 12-15 months, which is not part of the ELC score, was also significantly associated with cough (beta estimate =-0.05, CI=-0.09 - −0.02, p=0.005), fever (beta estimate =-0.19, CI=-0.28 - −0.11, p<0.001), and total illness weeks (beta estimate =-0.04, CI=-0.07 - −0.005, p=0.02), but not diarrhea/vomiting (p=0.73).

**Table 3:** Multivariable association* between cumulative illness in the first year of life, MSEL scores at 12-15 months, and presence of microcephaly or stunting at 12-15 months.

|  | # Total Illness Weeks |  | # Cough Weeks |  | # Fever Weeks |  | # Diarrhea/Vomiting Weeks |  |
| --- | --- | --- | --- | --- | --- | --- | --- | --- |
|  | Beta estimate<br>(95%<br>Confidence<br>Interval) | p-value | Beta estimate<br>(95%<br>Confidence<br>Interval) | p-value | Beta estimate<br>(95%<br>Confidence<br>Interval) | p-value | Beta estimate<br>(95% Confidence<br>Interval) | p-value |
| Early Learning Composite* | -0.04<br>(-0.11 – 0.03) | 0.27 | -0.08<br>(-0.16- 0.003) | 0.06 | -0.36<br>(-0.56 - -0.16) | 0.0004 | -0.03<br>(-0.09– 0.15) | 0.66 |
| Gross Motor* | -0.04<br>(-0.07 - -0.005) | 0.02 | -0.05<br>(-0.09 - -0.02) | 0.005 | -0.19<br>(-0.28 - -0.11) | <0.0001 | -0.009<br>(-0.47 – 0.06) | 0.73 |
| Fine Motor* | -0.01<br>(-0.04 – 0.02) | 0.42 | -0.02<br>(-0.05 – 0.01) | 0.20 | -0.03<br>(-0.05 – 0.0006) | 0.055 | 0.001<br>(-0.04 – 0.04) | 0.93 |
| Visual Receptive* | -0.01<br>(-0.04 – 0.01) | 0.30 | -0.03<br>(-0.06 - -0.002) | 0.03 | -0.07<br>(-0.14 - -0.005) | 0.03 | 0.02<br>(-0.02 – 0.06) | 0.42 |
| Expressive Language* | -0.008<br>(-0.05 – 0.03) | 0.65 | -0.02<br>(-0.06 – 0.02) | 0.31 | -0.11<br>(-0.20 - -0.01) | 0.03 | 0.02<br>(-0.03 – 0.08) | 0.44 |
| Receptive Language* | -0.009<br>(-0.03 – 0.009) | 0.33 | -0.01<br>(-0.03 – 0.008) | 0.24 | -0.08<br>(-0.13 - -0.03) | 0.002 | -0.01<br>(-0.05 – 0.02) | 0.37 |
|  | RR (95% CI) | p-value | RR (95% CI) | p-value | RR (95% CI) | p-value | RR (95% CI) | p-value |
| Stunting**,† | 1.00 (0.98-1.01) | 0.84 | 1.00 (0.98-1.02) | 0.79 | 0.8 (0.94-1.03) | 0.39 | 1.00 (0.98-1.02) | 0.95 |
| Microcephaly** | 0.99 (0.96-1.01) | 0.35 | 0.99 (0.96 – 1.02) | 0.66 | 0.95 (0.87-1.03) | 0.20 | 0.97 (0.93-1.02) | 0.29 |

Abbreviations: MSEL=Mullen Stages of Early Learning, SE=standard error, RR=relative risk
\*adjusted for age at last visit, sex, maternal height at end of study, breastfeeding at last visit, housing material, maternal education, food insecurity, and living in a community at high risk for infectious illness. Beta estimate reflects change in MSEL score per additional week of illness.
\*\*adjusted for sex, maternal height at end of study, housing material, and maternal education.
†Stunting defined as length-for-age <2 -SD on WHO growth curves 18.

Cumulative syndromic illness was distributed broadly across the study catchment area (**Figure 1 - online, Figure 2**). However, there were several communities that demonstrated a significantly greater number of weeks of illness than the overall mean (p<0.05). Living in a community at high risk for infectious illness was associated with neurodevelopmental outcome, but was not associated with the stunting or microcephaly outcomes; thus this ‘high risk for infectious illness’ community variable was included in the multivariable models of neurodevelopmental outcomes, but not the multivariable models of stunting or microcephaly outcomes (**Table 3**).

**Figure 1.**
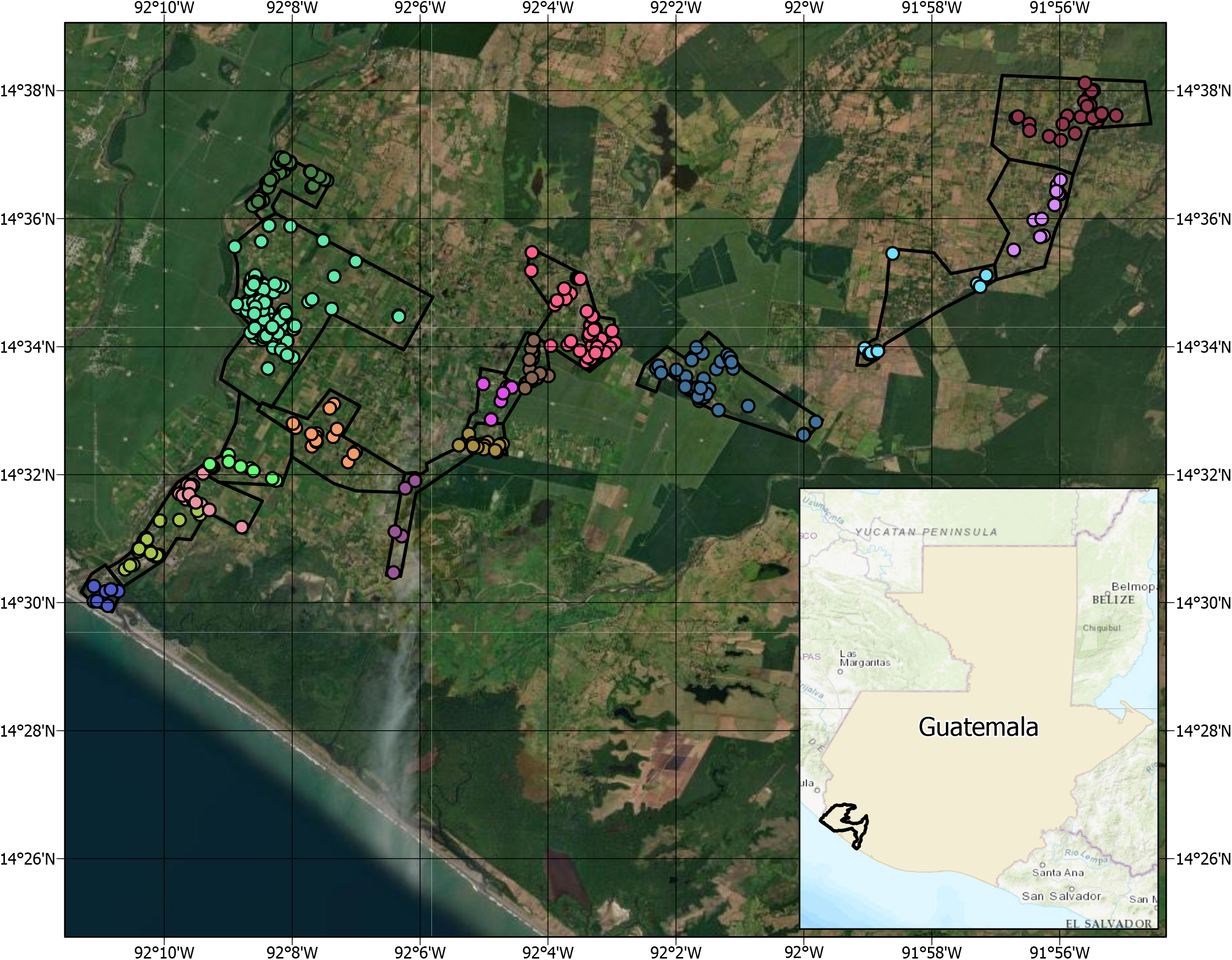
Households with Infants enrolled in Study and Grouped by Community in the Trifinio Region of SW Guatemala, 2017-2019 (online only) **Legend:** Household location of enrolled subjects, by community. Each dot represents a single household that completed the DMID 16-0057 study and was included in this secondary analysis. Each color represents a different community, which were compared to assess differences in cumulative illness burden by geolocation.

**Figure 2.**
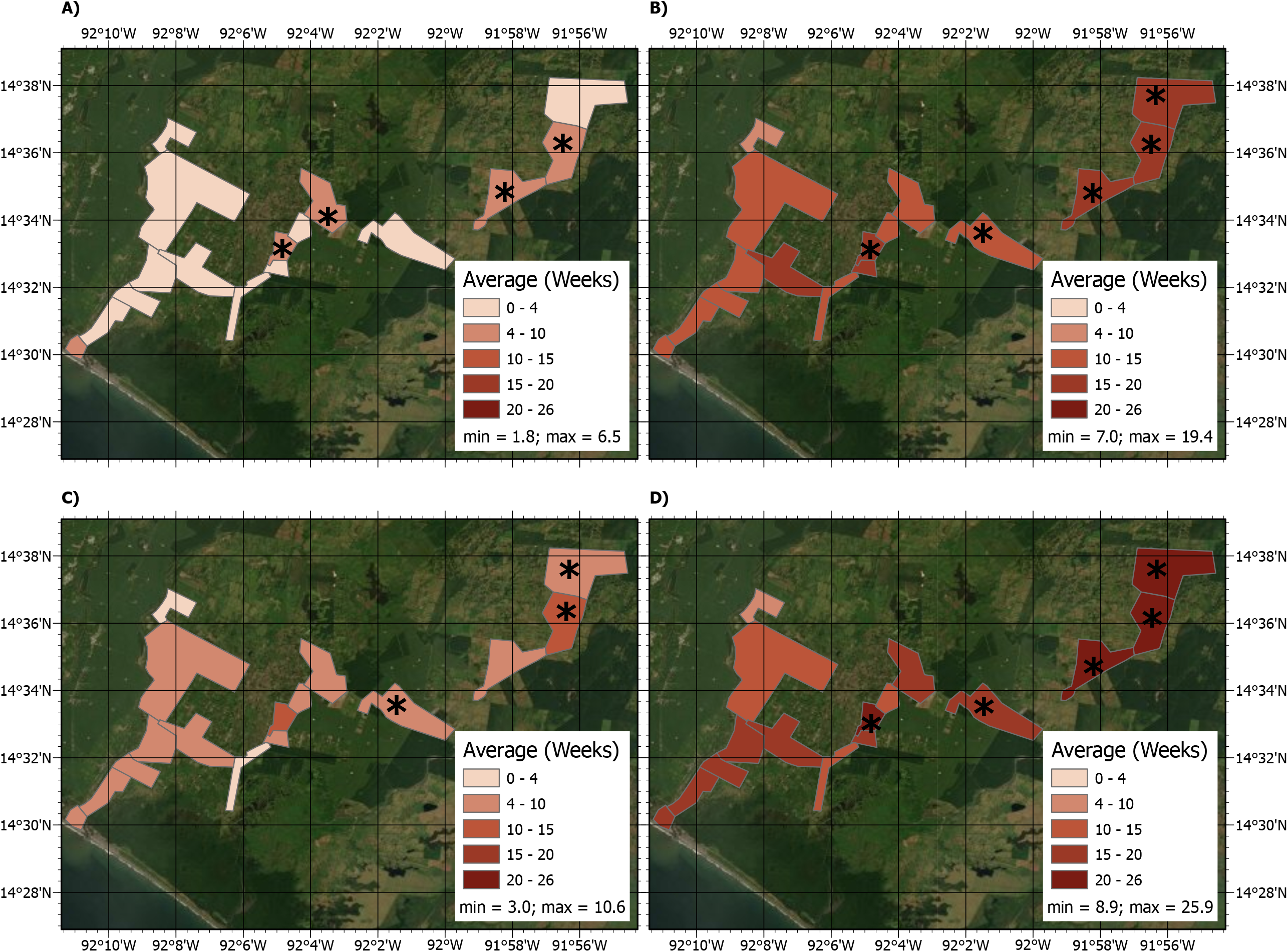
Cumulative Weeks of Syndromic Illness by Community among Infants in the Trifinio Region of SW Guatemala, 2017-2019. **Legend:** Mean weeks of cumulative illness by community for fever (Panel A), cough, (Panel B), diarrhea/vomiting (Panel C), or any illness (Panel D). Darker color indicates greater mean number of weeks of reported illness. Asterisks indicate communities that had significantly greater disease burden than the overall mean (Mann Whitney U Test: p<0.05).

### Stunting

Stunting was common, with 140 (32.6%) infants demonstrating a LAZ <-2 SD at 12-15 months. In univariate analysis, male sex (RR=2.03, CI=1.50 – 2.74, p<0.001), wood/aluminum housing material (RR=1.39, CI=1.04 – 1.86, p=0.02), lower maternal height (RR per −1 cm=0.97, CI=0.96 – 0.98, p<0.001), and lower maternal education (RR=0.82, CI=0.67 – 0.997, p=0.046) were significantly associated with stunting at 12-15 months (**Table X – online**). After adjusting for gender, house material, maternal height at end of study, and maternal education, there was no association between caregiver-reported cough, fever, or vomiting/diarrheal weeks of illness and stunting at 12-15 months (**Table 3**).

### Microcephaly

Microcephaly was also common, with 72 (16.7%) of infants demonstrating microcephaly (OFC <-2SD) at 12-15 months. Microcephaly was significantly associated with male sex (RR=1.64, CI=1.06 – 2.56, p=0.03) and marginally associated with maternal height at the end of the study (RR=0.97, CI=0.94 - - 1.002, p=0.06) on univariate analysis (**Table X - online**). After adjusting for sex, house material, maternal height at end of study, and maternal education, there was no association between caregiver-reported cough, fever, or vomiting/diarrheal weeks of illness and microcephaly at 12-15 months (**Table 3**).

## DISCUSSION

We found that cumulative weeks of febrile illness in the first year of life was significantly associated with worse neurodevelopmental outcome and cough trended towards significance, but cumulative weeks of syndromic illness did not have an association with stunting or microcephaly. Though some studies have identified harmful effects of cumulative illness in early childhood, including within Guatemala,^11^ few have evaluated neurodevelopment in a community-based longitudinal cohort using a rigorous, adapted, and validated measure by trained personnel in the assessment of neurodevelopmental outcomes.

Though several factors were associated with lower 12-15-month MSEL score, such as sex, maternal height, breastfeeding, housing type and maternal education, after adjusting for these variables, caregiver-reported weeks of fever was significantly associated with low MSEL, with a reduction in MSEL ELC raw score of 0.36 per week of reported illness (fever was also significantly associated with lower scores for all sub-domains). This corresponds with data from a Bangladeshi birth cohort, which found associations between febrile illness and systemic/enteric inflammatory cytokine profiles and decreased neurodevelopmental performance.^5-7, 9^ Reported weeks of cough did not achieve significance but also trended towards worse neurodevelopmental outcome.

Our findings provide additional insight into the complex pathways driving the neurodevelopment of young children in low resource settings. Accumulating evidence suggests that nutritional deficiency is only one of many contributors to poor neurodevelopmental outcome.^4, 8, 29, 30^ Importantly, as was the case with several variables, we did not find an association between cumulative weeks of illness and either stunting or microcephaly, supporting the hypothesis that cumulative febrile/respiratory illnesses may not negatively impact neurodevelopment through stunting or its biological consequences, at least during the short term. Accumulating data from multinational cohorts suggest that neurodevelopment may have a related, but distinct causal pathway from stunting.^9, 25-28, 31, 32^ Indeed, the aforementioned Bangladeshi cohort found that birth anthropometry and maternal weight were better predictors of growth, while clinical (fever) and biological markers of inflammation were more strongly associated with neurodevelopment.^5-7, 9^ Though not completely analogous to recurrent infections, mounting evidence suggests that human immunodeficiency virus (HIV)-infected, or even HIV-exposed, uninfected infants and children demonstrate neurodevelopment delays likely attributable to inflammation and immune dysregulation.^33, 34^ Putting our data into this context, we hypothesize that recurrent infections in our cohort negatively impact neurodevelopment in the first year of life, possibly through systemic inflammation, and not necessarily through stunting and its biological consequences. Further studies on the inflammatory phenotype of these infants may provide greater insight on the pathogenesis.

Interestingly, we did not find an association between diarrheal illness and stunting or neurodevelopment in our cohort. It is possible that within our population, the impact of diarrhea on growth would become more apparent in the second year of life. A smaller study in the Guatemalan highlands found that indigenous children (mean age 2.6 years) with >1 caregiver-reported illness (all-cause) per month did have increased risk of stunting, though they did not measure neurodevelopment.^35^ Other potential explanations are that pathogens in this community differ and/or that background rates of diarrhea are high enough in this community^36^ that our case definition (caregiver report of ≥3 loose stools/24 hours) did not adequately differentiate children with and without illness, or what is considered abnormal. Additionally, the type of inflammation and inflammatory pathways in diarrheal illness may be different than respiratory diseases, or they may be pathogen-specific to organisms less prevalent in this community.

Relying on caregiver report of illness was an overall limitation of the study, and our approach did not attempt to differentiate separate illnesses when children reported symptoms during sequential weeks, as this was likely to be inaccurate given the burden of overlapping childhood illnesses in this community. Weekly in-person visits reduced the risk of recall bias. In addition, we found that breastfeeding at 12-15 months was associated with decreased MSEL ECL at 12-15 months in both univariate and multivariable analyses. This is inconsistent with previously reported research on the protective factors associated with breastfeeding. We hypothesized that infants still breastfeeding at 12-15 months may have had greater food insecurity and/or lower socioeconomic status and therefore decreased neurodevelopment. Another hypothesis is that delayed neurodevelopment may lead to prolonged breastfeeding (reverse causality), and therefore breastfeeding was prolonged due to delayed neurodevelopment. Breastfed infants did have a non-significant increased risk of stunting. Other limitations include our lack of pathogen-specific testing (ZIKV infection is being evaluated separately) and self-reporting of some exposures (literacy, syndromic illness). Food insecurity was only collected at a single timepoint. However, food production in the community is generally not seasonal, and the prevalence of food insecurity reported in our cohort was similar to a 2016 (unpublished) survey of agricultural workers within the same community. Finally, we did not look at trajectories in growth outcomes (just absolute measures), and these should be included in further studies. Previous similar analyses have consistently found birth anthropometry to be the strongest predictor of anthropometry at later time points.

We did find some geographic heterogeneity in cumulative disease burden, with the three northeastern most communities showing the greatest burden. These findings suggest there may be some associated environmental risk factors (i.e., poverty, shared watershed, population density) that warrant further investigation.

In conclusion, cumulative, caregiver-reported febrile illness in a birth cohort of infants in rural Guatemala was associated with decreased neurodevelopmental outcomes at 12-15 months of age and respiratory illness was marginally associated with worse neurodevelopmental outcome. There was no association with syndromic illness and stunting or microcephaly. Further studies should evaluate for biomarkers of enteric and systemic inflammation, which may provide additional insight into the causal pathway between illness, inflammation, and neurodevelopmental outcome with longitudinal follow up beyond the first year of life.

## Data Availability

All data produced in the present study are available upon reasonable request to the authors

## Acknowledgements

The authors would like to thank Walla Dempsey, Gail Tauscher, Kay Tomashek (NIAID), and Wendy Keitel (Baylor College of Medicine) for their support of this project and their review of this manuscript. We also wish to thank the families of southwest Trifinio, Guatemala, who participated in this study and the research nurses and personnel from FUNSALUD who have worked on the parent study. Andrea Holliday, Chris Focht, Stephanie Pettibone, Nora Watson from EMMES. Mark Mulligan, Nadine Rouphael, Dean Kleinhenz, Michele McCullough, Erin Scherer, and Hannah Huston from Emory.

## Abbreviations and Acronyms

ELC: Early Learning Composite
LAZ: Length-for-age
MSEL: Mullen Scales of Early Learning
OFC: Occipital-frontal circumference
RR: Relative risk
SD: Standard deviations
SE: Standard error

---

**Supplemental Table 1:** Univariate association between predictor variables and presence of stunting (LAZ< - 2 SD) and microcephaly (OFC <-2SD) at 12-15 months.

| Variable | Stunting |  | Microcephaly |  |
| --- | --- | --- | --- | --- |
|  | RR (95% CI) | p-value | RR (95% CI) | p-value |
| # illnesses in first year of life | 1.00 (0.99-1.02) | 0.75 | 0.99 (0.96-1.02) | 0.41 |
| # cough illnesses in first year of life | 1.00 (0.98-1.02) | 0.81 | 0.99 (0.96-1.02) | 0.69 |
| # fever illnesses in first year of life | 0.98 (0.93-1.03) | 0.38 | 0.96 (0.87-1.03) | 0.19 |
| # diarrhea/vomiting illnesses in first year of life | 1.00 (0.98-1.03) | 0.73 | 0.97 (0.93-1.02) | 0.28 |
| Male sex | 2.03 (1.50-2.74) | <0.001 | 1.64 (1.06-2.56) | 0.03 |
| Lower maternal height (cm) at end of study | 0.97 (0.96-0.98) | <0.001 | 0.97 (0.94-1.002) | 0.06 |
| Maternal literacy | 0.78 (0.52-1.18) | 0.24 | 0.88 (0.44-1.78) | 0.73 |
| Maternal education | 0.82 (0.67-0.997) | 0.046 | 0.86 (0.63-1.17) | 0.34 |
| Paternal literacy | 0.70 (0.43-1.13) | 0.15 | 1.14 (0.39-3.33) | 0.82 |
| Paternal education | 0.84 (0.68-1.04) | 0.12 | 0.87 (0.62-1.23) | 0.43 |
| Housing material wood/aluminum/other | 1.39 (1.04-1.86) | 0.02 | 0.99 (0.60-1.65) | 0.98 |
| Caregiver-reported food insecurity | 1.09 (0.74-1.62) | 0.65 | 1.14 (0.63-2.08) | 0.67 |
| Child breastfeeding at 12-15 months | 1.25 (0.88-1.76) | 0.21 | 1.06 (0.64-1.73) | 0.83 |
| Food insecurity | 1.09 (0.74 – 1.62) | 0.65 | 1.12 (0.61 – 2.05) | 0.71 |
| Lives in community at high risk for infectious illness | 1.18 (1.88 – 1.56) | 0.27 | 0.81 (0.51 – 1.30) | 0.38 |

Abbreviations: LAZ=length-for-age, SD=standard deviation, OFC=occipital-frontal circumference,

## Notes

**Funding:** This project has been funded in whole or in part by the National Institute of Allergy and Infectious Diseases (NIAID), National Institutes of Health, Department of Health and Human Services. Research was supported by a NIAID Division of Microbiology and Infectious Diseases (DMID) Vaccine and Treatment Evaluation Unit (VTEU) award to Baylor College of Medicine (Contract No. HHSN27220130015I funding the DMID 16-0057 study, PIs: Munoz, Asturias) and EMMES (Contract No. 75N93021C00012). Daniel Olson is supported by K23AI143967 and CTSI Grant Number UL1 TR001082.

**Conflicts of interest:** none

### Competing Interest Statement

The authors have declared no competing interest.

### Funding Statement

This project has been funded in whole or in part by the National Institute of Allergy and Infectious Diseases (NIAID), National Institutes of Health, Department of Health and Human Services. Research was supported by a NIAID Division of Microbiology and Infectious Diseases (DMID) Vaccine and Treatment Evaluation Unit (VTEU) award to Baylor College of Medicine (Contract No. HHSN27220130015I funding the DMID 16-0057 study, PIs: Munoz, Asturias) and EMMES (Contract No. 75N93021C00012). Daniel Olson is supported by K23AI143967 and CTSI Grant Number UL1 TR001082.

